# The CoLab-score rapidly and efficiently excludes COVID-19 at the emergency department without need for SARS-CoV-2 testing: a multicenter case-control study

**DOI:** 10.1101/2024.01.30.24301996

**Authors:** Arjen-Kars Boer, Ruben Deneer, Maaike Maas, Heidi S. M. Ammerlaan, Roland H. H. van Balkom, Mathie P. G. Leers, Remy J.H. Martens, Madelon M. Buijs, Jos J. Kerremans, Muriël Messchaert, Jeroen D.E. van Suijlen, Natal A.W. van Riel, Volkher Scharnhorst

## Abstract

**Background:** Rapid identification of emergency department (ED) patients with a possible COVID-19 infection is needed. PCR-testing all ED patients is neither feasible nor effective in most centers, therefore a rapid, objective, low-cost screening tool to triage ED patients is necessary.

**Methods:** Results from all routine lab tests from ED patients at the Catharina Hospital were collected from July 2019 to July 2020 and used in a statistical model to obtain the CoLab-score. The score was validated temporally and externally in three independent centers.

**Results:** The CoLab-score consists of 10 routine lab results and can be used to safely rule-out a COVID-19 infection in more than one third of ED presentations with a negative predictive value of 0.997 (95% CI: 0.994 – 0.999).

**Conclusions:** The CoLab-score is a valuable tool to rule out COVID-19, guide PCR testing and is available to any center with access to routine laboratory tests.

## Introduction

A global pandemic caused by a novel virus like severe acute respiratory syndrome coronavirus 2 (SARS-CoV-2) disrupts society, the health system and hospitals in particular. Initially, little was known about the virus, its transmission, the diagnostic possibilities and treatment options. Combined with limited awareness in the community, the virus spread swiftly, resulting in exponential growth of infected patients. Interventions like social distancing and face masks subsequently resulted in a decline in prevalence of the virus. But after a low prevalence at the start of the summer, there was a revival of the virus in the fall, resulting in a second wave (“Coronavirus Disease (COVID-19) Situation Reports,” n.d.). Faced with novel mutations in SARS-CoV-2, in many countries a third wave is very likely to occur. In all phases of the outbreak, emergency departments (EDs) faced different diagnostic challenges. Often, the capacity of the SARS-CoV-2 RNA PCR testing facility was limited and patient care was delayed due to the long turnaround time of centralized PCR facilities. Shortly after the first wave, rapid SARS-CoV-2 tests became available, potentially reducing the diagnostic phase to less than 1 hour.

Prediction models that reliably identify ED patients eligible for PCR testing, or safely rule-out patients without coronavirus disease 2019 (COVID-19), are desirable. Moreover, if only routine laboratory results are required as input, a model could potentially reduce diagnostic costs and accelerate triaging of patients. In the era of algorithms and artificial intelligence, it is very appealing to develop predictive or diagnostic tools based on information readily available. In fact, models have been developed to identify SARS-CoV-2-positive patients (Brinati et al., 2020; Fink et al., 2020; Giamello et al., 2020; Joshi et al., 2020; Kurstjens et al., 2020; Plante et al., 2020; Qin et al., 2020; Tordjman et al., 2020), to describe the severity of COVID-19 symptoms or to predict deterioration or ultimately admission at the intensive care unit (Linssen et al., 2020; Wang et al., 2020; Zeng et al., 2021; Zhou et al., 2020). For an extensive overview and critical appraisal of models developed for COVID-19 we refer to the living systematic review by Wynants et. al (Wynants et al., 2020). However, only few models found their way into routine care at the ED (Albahri et al., 2020; Hooli and King, 2020). Early models were based on relatively small sample sizes, were hampered by selection bias or required features that could not be acquired automatically. Furthermore, models are easily over-fitted by selecting too many features, resulting in models that could not be used outside the strict inclusion criteria (Albahri et al., 2020; Hooli and King, 2020; Wynants et al., 2020). Models developed on a population of PCR-tested patients, cannot be used to guide clinicians to decide for PCR testing, as the clinical considerations to request PCR testing are not taken into account. In addition, when volume and speed are important, also composite models requiring data from other diagnostic tools like a CT scan (COVID-19 CO-RADS classification) (Lessmann et al., 2021) or X-ray (Kurstjens et al., 2020) are hard to implement in routine care.

In this study we report on the development and validation of a prediction model that, based on routine laboratory tests, estimates the risks of a patient presenting at the ED for testing positive for COVID-19. The model can be used during high and low prevalence (“waves”) of COVID-19. Since the model was successfully validated externally, it can be used clinically to reduce the number of PCR tests, exclude COVID-19 in patients and identify patients with a high risk of being SARS-CoV-2 positive, even if the patients are asymptomatic.

## Methods

### Study design

Routine laboratory results combined with age and gender from all patient presenting at the emergency department (ED) of the Catharina Hospital Eindhoven from July 2019 to July 2020 were combined with SARS-CoV-2 PCR test results in a development dataset. A model that could predict the presence of a COVID-19 infection was fit to this dataset. Performance of the model was assessed by 1) internal validation using bootstrapping, 2) temporal validation by using data from all patients presenting at the ED of the Catharina Hospital from July 2020 to November 2020, and 3) external validation by using data from the ED of three other hospitals in the Netherlands. The study was reviewed by the Medical research Ethics Committees United (MEC-U) under study number W20.071, which confirmed that the Medical Research Involving Human Subjects Act (In Dutch: WMO) does not apply to this study. The study was thereafter reviewed and approved by the internal hospital review board.

### Development dataset

All ED patient presentations of the Catharina Hospital Eindhoven from July 2019 to July 2020 were included in the development dataset, provided that routine laboratory testing had been requested by the attending physician. The routine laboratory panel at the ED consists of 28 laboratory tests. However, in some cases not all tests in the routine panel were requested or one or more quantitative results were not available due to analytical interference (hemolysis, lipemia or icterus). Presentations with missing values in the routine panel were excluded. Presentations with one or more extreme lab results (> 10 times standard deviation) were also excluded. After the first case of COVID-19 in the Netherlands (28 February 2020) all patients with symptoms of COVID-19 (either fever and/or respiratory symptoms) were subjected to nasopharyngeal PCR testing for SARS-CoV-2 RNA. PCR testing was performed by commercial tests that were approved by the Dutch national institute of public health (RIVM). If a patient had a positive PCR result in the past, subsequent presentations were excluded as re-presentations might be clinically different from de novo presentations.

The ED routine lab panel results were merged with SARS-CoV-2 PCR results if the underlying nose or throat swap had been taken ≤ 1 day prior, or ≤ 1 week after initial blood withdrawal at the ED. If multiple PCR tests were performed in this window and at least one PCR test was positive, the presentation was labelled “*PCR-positive*”. If all PCR test results in the time window were negative, the presentation was labelled as “*PCR-negative*”. If no PCR tests were performed in the time window but the presentation occurred after the first case of COVID-19 in the Netherlands, the presentation was labelled as “*Asymptomatic*”. All presentations before the first case in our center were labelled as “*Pre-COVID-19*”.

### Laboratory tests

The routine laboratory panel consisted of hemocytometric and chemical analyses. In the Catharina hospital, the hemocytometric tests were performed on Sysmex XN-10 instruments (Sysmex Corp., Kobe, Japan) and consisted of hemoglobin, hematocrit, erythrocytes, mean corpuscular volume (MCV), mean cellular hemoglobin (MCH), mean cellular hemoglobin concentration (MCHC), thrombocytes, leukocytes, neutrophils, eosinophils, basophils, lymphocytes and monocytes. The chemical analyses were performed on a Cobas 8000 Pro (Roche Dx, Basel, Switzerland) instrument and consisted of glucose, total bilirubin, aspartate aminotransferase (ASAT), alanine aminotransferase (ALAT), lactate dehydrogenase (LD), creatine kinase (CK), alkaline phosphatase (ALP), gamma-glutamyltransferase (gGT), blood urea nitrogen (BUN), creatinine, CKD-epi estimated glomerular filtration rate (eGFR), potassium, sodium, chloride, albumin (bromocresol green) and C-reactive protein (CRP). These results were combined with age and gender.

### Modelling

All data were processed and analyzed in R version 4.0.3 (R Core Team, 2020). Laboratory results, combined with age and gender were used as covariates in the model. Cases were ED presentations labelled as “*PCR-positive*”, controls as “*PCR-negative*”, “*Asymptomatic*” or “*Pre-COVID-19*”. To minimize additional missing data, all non-numeric results at the extremes of the measuring range, were converted to numeric results by removing the “<” and “>” signs. For eGFR (CKD-epi) and CRP this caused a discontinuous distribution. Therefore, instead of random sampling for eGFR (CKD-epi) and CRP the raw precursor value was used instead of >90 ml/min/m^2^ and <6 mg/L, respectively. Considering that laboratory results of bilirubin, ASAT, ALAT, LD, CK, AF and gGT can have heavy (right) tailed distributions, which in turn impacts model predictions, these variables were transformed logarithmically.

To achieve predictive accuracy, limit overfitting and emphasize sparsity, adaptive lasso regression was chosen (Tibshirani, 1996; Zou, 2006). Prior to model fitting, covariates were scaled to zero mean and unit variance, after model fitting coefficients were unscaled to obtain regression coefficients on the original scale. In adaptive lasso, weights are applied to each of the covariates present in the lasso constraint, the weight vector has to be calculated before the adaptive lasso regression is performed. Due to multicollinearity between laboratory tests in the routine lab panel, weights in the adaptive lasso were based on ridge regression estimates (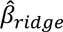) as recommended by Zou (Zou, 2006). To obtain 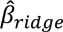 the optimal penalty (λ) for the ridge regression was chosen using 10 fold cross-validation (CV) with area under the ROC curve (AUC) as the loss function. The λ corresponding to the maximum AUC was selected to obtain 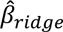. The weight vector (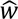) was calculated by 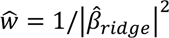. This weight vector was then used to fit an adaptive lasso regression where λ was chosen by the criterion ±1 SE of the maximum AUC. All models were fitted using the glmnet-package (Friedman et al., 2010).

Since this is a retrospective case-control study, the sample prevalence may not reflect the true/current prevalence, as is the case with COVID-19. Therefore the intercept term in the model must be adjusted according to the actual prevalence (if known, or estimated) to obtain well-calibrated probabilities (King and Zeng, 2001).

### CoLab-score

Since manually updating the intercept term is not straightforward to implement in clinical practice, we developed the CoLab-score. The CoLab-score is a categorization of the linear predictor into a CoLab-score ranging from 0 to 5, where score 0 reflects low risk for COVID-19 and score 5 reflects high risk. The CoLab-score is based on a fixed number needed to test of 15, this implies that one is willing to test 15 patients to find one positive. A number needed to test of 15 corresponds to a threshold probability of 1/15 = 0.067, on the linear predictor (or log-odds) scale, this corresponds to a threshold of logit(0.067) = −2.639. This threshold is correct when the sample prevalence matches the actual prevalence. In reality the prevalence of COVID-19 in the ED varies, therefore thresholds are calculated for prevalence cut-points of 1%, 2%, 5%, 10% and 40% using the intercept adjustment formula by King (King and Zeng, 2001). The intervals obtained through these breaks correspond to CoLab-scores 5 to 0, respectively. The CoLab-score can thus be used as a “prevalence-dependent binary classification” (either positive or negative). If COVID-19 prevalence at the ED in the previous week is <1%, only CoLab-score 5 should be classified as positive and CoLab-score 0 till 4 as negative. If prevalence is 1% – 2%, CoLab-score 4 and 5 should be classified as positive and 1 – 3 negative. Similarly, with a prevalence of 2 – 5% the split is between CoLab-score 2 and 3 and with prevalence of 5 – 10% between CoLab-score 1 – 2. If the prevalence is higher than 10% only CoLab-score 0 is classified as negative. Using the CoLab-score in this fashion, aims to preserve a number need to test of 15.

### Internal validation

To assess model performance while taking overfitting into account, bootstrapping was performed. 1000 bootstrap samples were generated from the original data. On each bootstrap sample, the full model fitting procedure and CoLab-score conversion were performed. Optimism adjusted performance measures of the CoLab-score were obtained by applying the 0.632 bootstrap rule to the in-sample and out-of-bag-sample performance (Efron, 1983). Performance measures included, AUC, sensitivity, specificity, positive predictive value (PPV) and negative predictive value (NPV) of each CoLab-score. The pROC-package was used to calculate performance measures (Robin et al., 2011). The performance of the prevalence dependent binary CoLab-classification was evaluated in terms of overall sensitivity and specificity, and potential reduction in the number of PCR-tests. Although the full inclusion period from July 2019 to July 2020 was used for model fitting, the performance was evaluated on the period starting from the first COVID-19 infection (24^th^ of February 2020) to July 2020. This was done to obtain performance measures that would reflect real world performance during COVID-19 waves.

### Temporal validation

For temporal validation, results from our center were prospectively analyzed from July 2020 to November 2020. During this period, a second wave of COVID-19 infections developed. In this period there was also more widespread external PCR testing by municipal health services (In Dutch: GGD). Since the results of external conducted PCR tests were not available to our study, the outcome in the temporal validation cohort was chosen as a composite of the hospital registration of a confirmed COVID-19 infection or at least one positive PCR test results in the hospital (with the same time window as described above). The temporal validation consisted of assessing the AUC, sensitivity, specificity, PPV and NPV of each CoLab-score threshold. Calibration was assessed graphically by converting the CoLab-linear predictor to the predicted probability (through the inverse logit function) and plotting the proportion of observed outcomes versus expected probabilities. The rms-package was used to generate calibration plots (Harrell Jr, 2021).

### External validation

For the external validation, several centers in the Netherlands were approached and asked if they collected the required panel of laboratory tests and had access to SARS-CoV-2 PCR test results. Seven centers responded and the three centers with the least inclusion bias (i.e. measuring routinely all laboratory tests underlying the CoLab-score for large unselected patient groups) were Gelre Hospitals (center 1), Atalmedial Diagnostic Centers, location Alrijne Hospital Leiderdorp (center 2) and Zuyderland Medical Center (center 3). The hematological parameters were measured with Sysmex XN10/XN20 (center 1), CELL-DYN-Sapphire (Abbott Laboratories) (center 2) and Sysmex XN10 instruments (center 3). The clinical chemistry parameters were measured with Architect c14100/c160000 (Abbott Laboratories) (center 1), Architect ci4100 (Abbott Laboratories) (center 2) and Cobas 8000 instruments (Roche Dx) (center 3). The external validation was similar to the temporal validation and consisted of assessing the AUC sensitivity, specificity, PPV and NPV of each CoLab-score threshold. Calibration was assessed graphically analogous to the temporal validation dataset. In addition, the distributions of the variables in the CoLab-scores was plotted for all control (i.e. negative) patients to determine if there were any systematic differences that could cause bias in the CoLab-score predictions.

## Results

### Development dataset

For model building, 12.879 emergency department (ED) presentations of 10.327 patients from July 2019 to July 2020 were included. After excluding cases with an incomplete lab panel, patient presentations that occurred after a positive PCR test in the past (re-presentations) and presentations with extreme values (>10 times standard deviation) in any of the lab results, 10.417 presentations of 8610 patients remained (**Figure 1A**).

**Figure 1:**
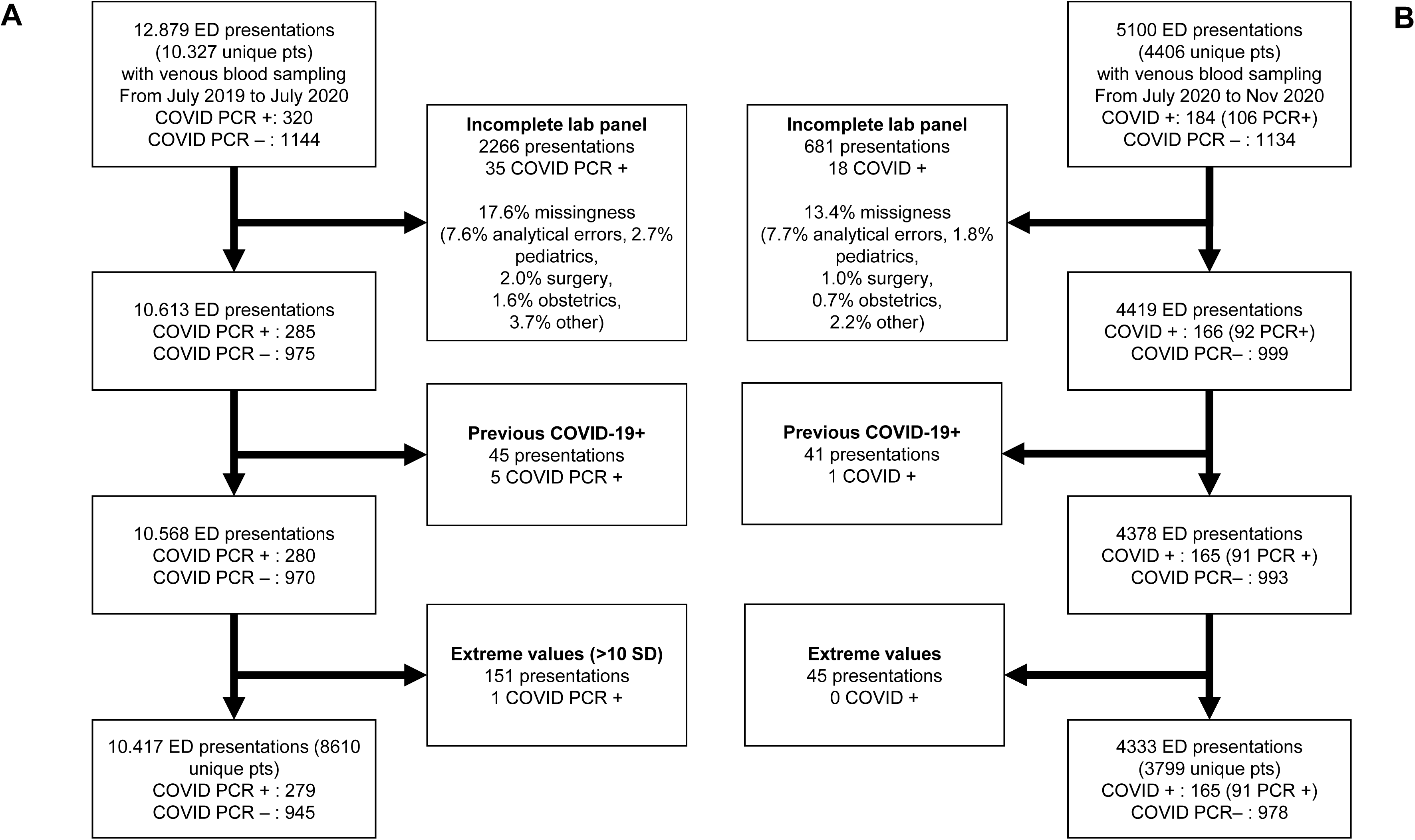
Inclusion flow of patients in the development (A) and temporal validation (B) dataset. All patient admissions with routine venous blood sampling at the emergency department (ED) were included. For the development dataset, completeness of the lab panel was assessed for all the 29 laboratory tests (see Table 1), for the temporal validation dataset this was only necessary for 10 laboratory tests (see Table 2). The major causes of missingness are described in the text. In the development dataset, presentations with extreme values (>10 SD) were excluded. The same limits were applied to the temporal validation dataset (see Table 2 for limits).

**Table 1:**
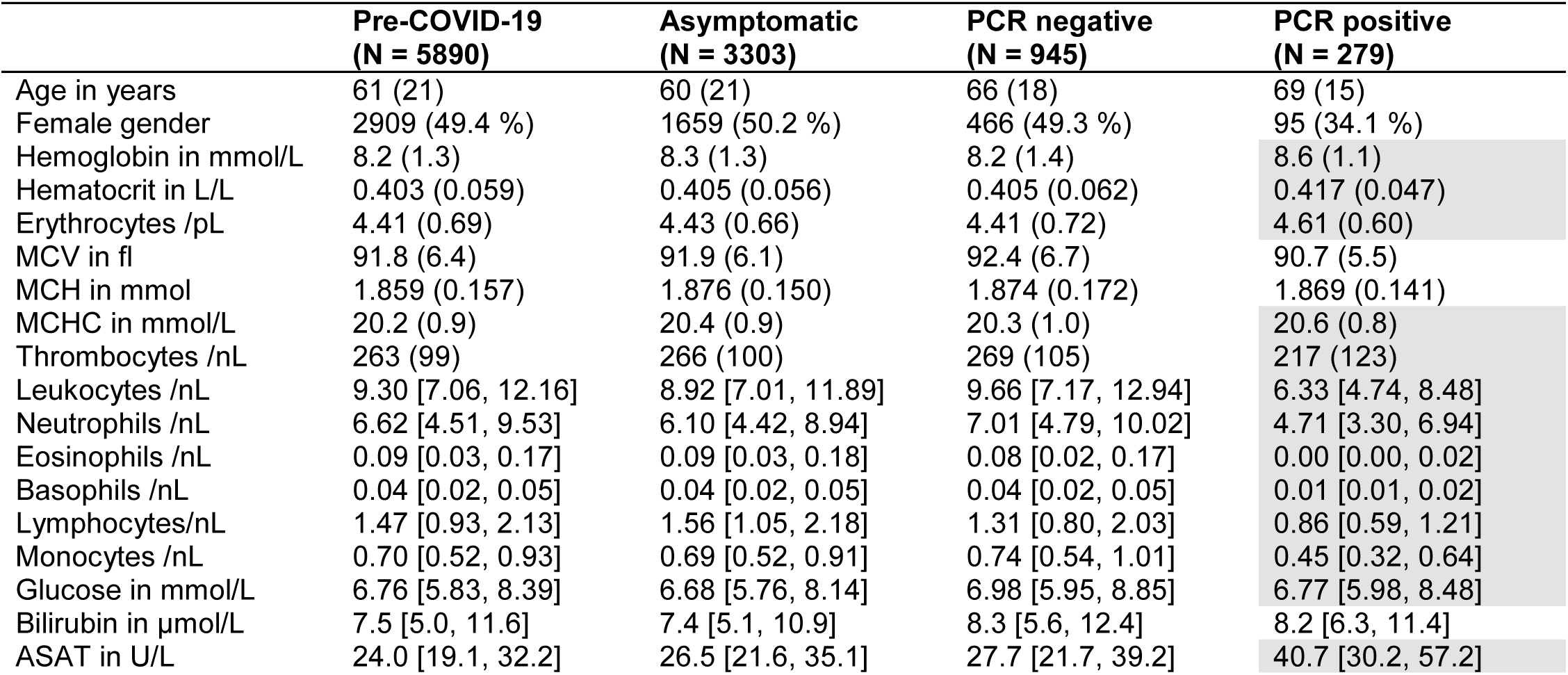
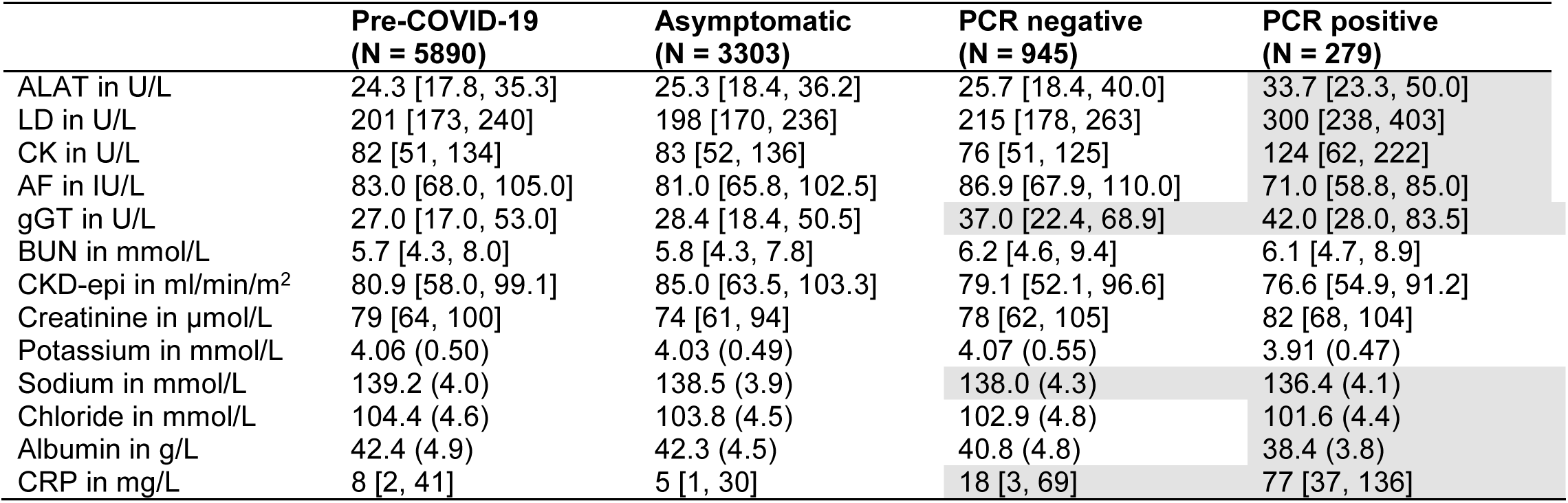
Descriptive statistics of development dataset and laboratory concentrations. Shown are the laboratory tests routinely requested at ED presentation and their mean/median results (in the development dataset) for the presentations before the first COVID-19 patient in the Netherlands (“Pre-COVID-19”), presentations thereafter that were not tested for COVID-19 (“Asymptomatic”), tested negatively (“PCR negative”) and tested positive (“PCR positive”). For results with normal distributions, the mean value and standard deviation (in round brackets) are shown. For results that have skewed or heavy tailed distributions, the median value and the interquartile range is shown [in squared brackets]. Dark grey marked figures indicate a clinically relevant difference from the Pre-COVID-19 category (based on the total allowable error (Ricós et al., 1999)).

|  | Pre-COVID-19<br>(N = 5890) | Asymptomatic<br>(N = 3303) | PCR negative<br>(N = 945) | PCR positive<br>(N = 279) |
| --- | --- | --- | --- | --- |
| Age in years | 61 (21) | 60 (21) | 66 (18) | 69 (15) |
| Female gender | 2909 (49.4 %) | 1659 (50.2 %) | 466 (49.3 %) | 95 (34.1 %) |
| Hemoglobin in mmol/L | 8.2 (1.3) | 8.3 (1.3) | 8.2 (1.4) | 8.6 (1.1) |
| Hematocrit in L/L | 0.403 (0.059) | 0.405 (0.056) | 0.405 (0.062) | 0.417 (0.047) |
| Erythrocytes /pL | 4.41 (0.69) | 4.43 (0.66) | 4.41 (0.72) | 4.61 (0.60) |
| MCV in fl | 91.8 (6.4) | 91.9 (6.1) | 92.4 (6.7) | 90.7 (5.5) |
| MCH in mmol | 1.859 (0.157) | 1.876 (0.150) | 1.874 (0.172) | 1.869 (0.141) |
| MCHC in mmol/L | 20.2 (0.9) | 20.4 (0.9) | 20.3 (1.0) | 20.6 (0.8) |
| Thrombocytes /nL | 263 (99) | 266 (100) | 269 (105) | 217 (123) |
| Leukocytes /nL | 9.30 [7.06, 12.16] | 8.92 [7.01, 11.89] | 9.66 [7.17, 12.94] | 6.33 [4.74, 8.48] |
| Neutrophils /nL | 6.62 [4.51, 9.53] | 6.10 [4.42, 8.94] | 7.01 [4.79, 10.02] | 4.71 [3.30, 6.94] |
| Eosinophils /nL | 0.09 [0.03, 0.17] | 0.09 [0.03, 0.18] | 0.08 [0.02, 0.17] | 0.00 [0.00, 0.02] |
| Basophils /nL | 0.04 [0.02, 0.05] | 0.04 [0.02, 0.05] | 0.04 [0.02, 0.05] | 0.01 [0.01, 0.02] |
| Lymphocytes/nL | 1.47 [0.93, 2.13] | 1.56 [1.05, 2.18] | 1.31 [0.80, 2.03] | 0.86 [0.59, 1.21] |
| Monocytes /nL | 0.70 [0.52, 0.93] | 0.69 [0.52, 0.91] | 0.74 [0.54, 1.01] | 0.45 [0.32, 0.64] |
| Glucose in mmol/L | 6.76 [5.83, 8.39] | 6.68 [5.76, 8.14] | 6.98 [5.95, 8.85] | 6.77 [5.98, 8.48] |
| Bilirubin in μmol/L | 7.5 [5.0, 11.6] | 7.4 [5.1, 10.9] | 8.3 [5.6, 12.4] | 8.2 [6.3, 11.4] |
| ASAT in U/L | 24.0 [19.1, 32.2] | 26.5 [21.6, 35.1] | 27.7 [21.7, 39.2] | 40.7 [30.2, 57.2] |

**Table 1: Descriptive statistics of development dataset and laboratory concentrations.**
|  | **Pre-COVID-19**<br>**(N = 5890)** | **Asymptomatic**<br>**(N = 3303)** | **PCR negative**<br>**(N = 945)** | **PCR positive**<br>**(N = 279)** |
| --- | --- | --- | --- | --- |
| ALAT in U/L | 24.3 [17.8, 35.3] | 25.3 [18.4, 36.2] | 25.7 [18.4, 40.0] | 33.7 [23.3, 50.0] |
| LD in U/L | 201 [173, 240] | 198 [170, 236] | 215 [178, 263] | 300 [238, 403] |
| CK in U/L | 82 [51, 134] | 83 [52, 136] | 76 [51, 125] | 124 [62, 222] |
| AF in IU/L | 83.0 [68.0, 105.0] | 81.0 [65.8, 102.5] | 86.9 [67.9, 110.0] | 71.0 [58.8, 85.0] |
| gGT in U/L | 27.0 [17.0, 53.0] | 28.4 [18.4, 50.5] | 37.0 [22.4, 68.9] | 42.0 [28.0, 83.5] |
| BUN in mmol/L | 5.7 [4.3, 8.0] | 5.8 [4.3, 7.8] | 6.2 [4.6, 9.4] | 6.1 [4.7, 8.9] |
| CKD-epi in ml/min/m<sup>2</sup> | 80.9 [58.0, 99.1] | 85.0 [63.5, 103.3] | 79.1 [52.1, 96.6] | 76.6 [54.9, 91.2] |
| Creatinine in µmol/L | 79 [64, 100] | 74 [61, 94] | 78 [62, 105] | 82 [68, 104] |
| Potassium in mmol/L | 4.06 (0.50) | 4.03 (0.49) | 4.07 (0.55) | 3.91 (0.47) |
| Sodium in mmol/L | 139.2 (4.0) | 138.5 (3.9) | 138.0 (4.3) | 136.4 (4.1) |
| Chloride in mmol/L | 104.4 (4.6) | 103.8 (4.5) | 102.9 (4.8) | 101.6 (4.4) |
| Albumin in g/L | 42.4 (4.9) | 42.3 (4.5) | 40.8 (4.8) | 38.4 (3.8) |
| CRP in mg/L | 8 [2, 41] | 5 [1, 30] | 18 [3, 69] | 77 [37, 136] |

**Table 2:** Calculation of the CoLab-linear predictor (LP). The CoLab-linear predictor (LP) is calculated by summing the intercept and the products of the 11 variables with their corresponding coefficients (β’s). CoLab-LP = – 6.885 + [erythrocytes] × 0.9379 – [leukocytes] × 0.1298 – [eosinophils] × 6.834 – [basophils] × 47.7 – log10([bilirubin]) × 1.142 + log10([LD]) × 5.369 – log10([AF]) × 3.114 + log10([gGT]) × 0.3605 – [albumin] × 0.1156 + [CRP] × 0.02560 + [age] × 0.002275. The LP can be converted into a CoLab-score (see Figure 2) or into a probability if the prevalence is known or estimated (see details in Appendix A). The CoLab-score is not valid if any of the variables exceed the limits in the third column.

| Variable | $\beta$ | Exclusion limit | Relative importance |
| --- | --- | --- | --- |
| Intercept | -6.885 |  | - |
| Erythrocytes /pL | 0.9379 | Erythrocytes < 2.9 /pL | 52 % |
| Leukocytes /nL | -0.1298 |  | 46 % |
| Eosinophils /nL | -6.834 |  | 86 % |
| Basophils /nL | -47.70 | Basophils >0.33 /nL | 100 % |
| log<sub>10</sub> of Bilirubin in μmol/L | -1.142 | Bilirubin >169 μmol/L | 26 % |
| log<sub>10</sub> of LD in U/L | 5.369 | LD >1564 U/L | 58 % |
| log<sub>10</sub> of AF in IU/L | -3.114 | AF >1000 IU/L | 45 % |
| log<sub>10</sub> of gGT in U/L | 0.3605 | gGT >1611 U/L | 11 % |
| Albumin in g/L | -0.1156 |  | 45 % |
| CRP in mg/L | 0.002560 |  | 15 % |
| Age in years | 0.002275 |  | 4 % |

Descriptive statistics of ED presentations are shown in **Table 1**. Presentations were stratified into four categories: “*Pre-COVID-19*” consists of all ED presentations before the first case of COVID-19 in our center (N = 5890). This period also includes a winter flu-season. The category “A*symptomatic*” consists of ED presentations (N = 3303) after the first case of COVID-19, but who were not subjected to PCR testing. Finally, presentations tested for SARS-CoV-2 were categorized depending on the result as “*PCR negative*” (N= 945) or “*PCR positive*” (N= 279). In the latter category, 91% (95% CI: 88 to 94%) of the cases were tested positive in their first PCR. The remaining 24 patients were positive in their second (N = 18), third (N = 5) or fourth (N = 1) PCR.

**Table 1** shows the mean/median concentrations of the ED laboratory tests for each category. Concentrations that clinically differed from the Pre-COVID-19 category are marked in grey. As expected, patients tested positive for SARS-CoV-2 showed multiple clinically relevant differences. Interestingly, some differences (like increased CRP and gGT) were – although to a lesser extent – also present in the COVID-19 negative-tested patient group.

### CoLab-score

The model obtained through adaptive lasso regression contained eleven variables, which are depicted with their regression coefficients (weights) in **Table 2**. A larger β-coefficient does not imply that a variable is more important in predicting the odds of testing positive for SARS-CoV-2, since the coefficients are unscaled after model fitting and variables are on different scales. Therefore, the relative importance is calculated based on scaled coefficients. The absolute basophil count has the highest relative weight, followed by eosinophil count.

As shown in **Figure 2**, the linear predictor clearly discriminates between COVID-19 and non-COVID-19. Note that in this figure, the area under both curves is equal since these are probability density functions, in reality the weekly COVID-19 prevalence at the ED varies from <1% to 36% and hence the number of presentations in the PCR positive group. Therefore, the linear predictor was converted to CoLab-scores 0 – 5 (**Figure 2**). Subsequently, the actual prevalence of the previous week was used to obtain a threshold above which patients should be tested, this is hereafter referred to as the “prevalence-dependent binary classification”. CoLab-scores above the prevalence-dependent cut-off value, are classified as “positive” and warrant PCR testing, while CoLab-scores equal to or below the cut-off value (= ”negative”) discourage PCR testing. If the prevalence is not known, the CoLab-score can be used as-is, where patients with a CoLab-score of 0 can be considered non-COVID-19 and patients with a score 5 as highly suspicious for COVID-19.

**Figure 2:**
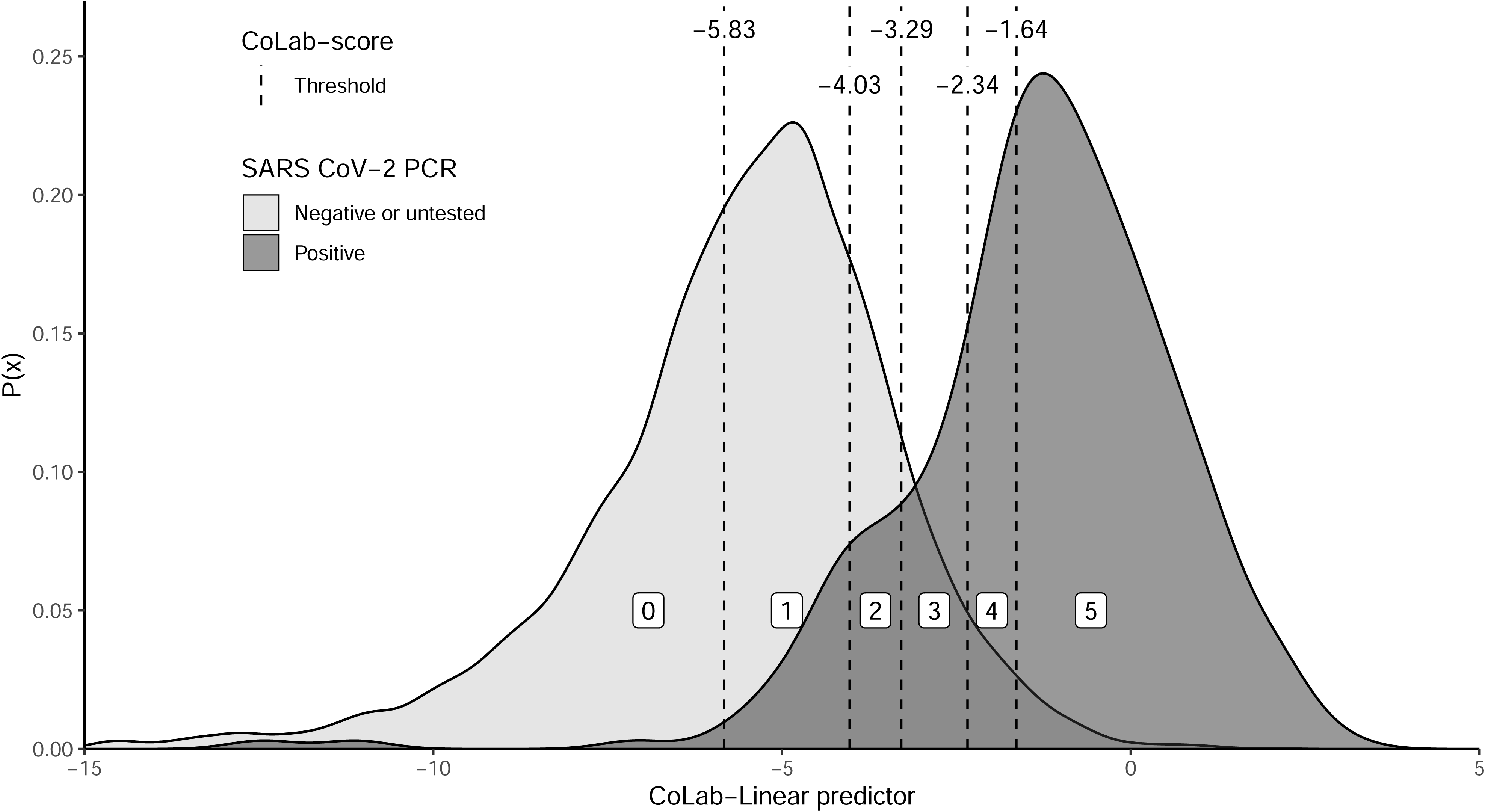
Probability density plot of the CoLab-linear predictor. The probability density plots for COVID (dark grey) and non-COVID patients (light grey) are plotted against the linear predictor (see table 2). The CoLab-score cut-offs (–5.83, –4.02, –3.29, –2.34 and –1.64) are depicted with vertical dashed lines. The white-boxed numbers (between the cut-offs) represent the corresponding CoLab-score. Note that while the area under both curves is identical (since these are probability density functions), in absolute numbers the “negative or untested”-group is about 36 times larger than the PCR positive group.

### Internal validation

The model was validated on the period starting from the first COVID-19 infection to July 2020, in this period the mean prevalence was 7.2%. The AUC of the CoLab-score is 0.930 (95% CI: 0.909 to 0.945). A CoLab-score of 0 has a negative predictive value (NPV) of 0.997 (95% CI: 0.994 to 0.999, **Table 3**), one third (38.4%, 95% CI: 26.4 to 48.4%) of all ED presentations were assigned this score and can therefore be safely excluded. Conversely, 6.2% (95% CI: 6.3 to 7.2%) of the ED patients have a CoLab-score 5. Given the high positive predictive value (PPV) of this category (0.682, 95% CI: 0.622 to 0.740), subsequent PCR testing is advised.

**Table 3:** Diagnostic performance CoLab-score in the development dataset. The development dataset was internally validation for the period March 2020 – July 2020 (N = 4.527). Sensitivities, specificities, positive predictive values (PPV), negative predictive values (NPV) and fraction of patients (%) are shown for fixed cut-offs (CoLab-score 0 till ≤ 4) and the prevalence dependent binary CoLab-classification (Pos/Neg). The numbers in round brackets represent the 95% confidence intervals. The first column defines the threshold above which CoLab-score a patient is considered positive.

| CoLab Cut-off | Sensitivity | Specificity | PPV | NPV | % |
| --- | --- | --- | --- | --- | --- |
| 0 | 0.984 (0.970 - 0.991) | 0.411 (0.285 - 0.520) | 0.115 (0.0932 - 0.141) | 0.997 (0.994 - 0.999) | 38.4 (26.4 - 48.4) |
| ≤ 1 | 0.909 (0.886 - 0.943) | 0.793 (0.744 - 0.826) | 0.255 (0.207 - 0.299) | 0.991 (0.989 - 0.995) | 74.4 (69.4 - 77.4) |
| ≤ 2 | 0.859 (0.811 - 0.889) | 0.887 (0.866 - 0.901) | 0.371 (0.317 - 0.414) | 0.988 (0.983 - 0.991) | 83.2 (82.2 - 85.2) |
| ≤ 3 | 0.750 (0.700 - 0.810) | 0.953 (0.944 - 0.959) | 0.551 (0.494 - 0.601) | 0.980 (0.975 - 0.985) | 90.1 (89.1 - 91.1) |
| ≤ 4 | 0.608 (0.522 - 0.685) | 0.978 (0.973 - 0.983) | 0.682 (0.622 - 0.740) | 0.970 (0.962 - 0.977) | 93.8 (92.8 - 93.8) |
| Pos/Neg | 0.923 (0.879 - 0.944) | 0.816 (0.756 - 0.842) | 0.310 (0.246 - 0.352) | 0.992 (0.987 - 0.994) | - |

For CoLab-scores 1 - 4, the threshold to guide PCR testing is dependent on COVID-19 prevalence. Therefore, the CoLab-score was dichotomized as positive or negative, based on the prevalence of the previous week. As shown in **Table 3**, the sensitivity of this binary CoLab-classification (0.923, 95% CI: 0.879 to 0.944) is similar to singular PCR testing (0.91, 95% CI: 0.88 to 0.94), and has a NPV of 0.992 (95% CI: 0.987 to 0.994). Therefore, the CoLab-score and the prevalence dependent binary classification could be used to guide PCR testing when rapid PCR testing is not available or test capacity is limited. Using the CoLab-score retrospectively on the development dataset to guide PCR testing, 92.3% (95% CI: 87.9 to 94.4%) of the COVID-19 patients could have been identified with 30.5% (95% CI: 14.5% to 37.6%) fewer PCR test. The largest reductions are seen in periods with low or rapidly decreasing prevalence, when more than half of the PCR tests may be superfluous (**Figure 3**).

**Figure 3:**
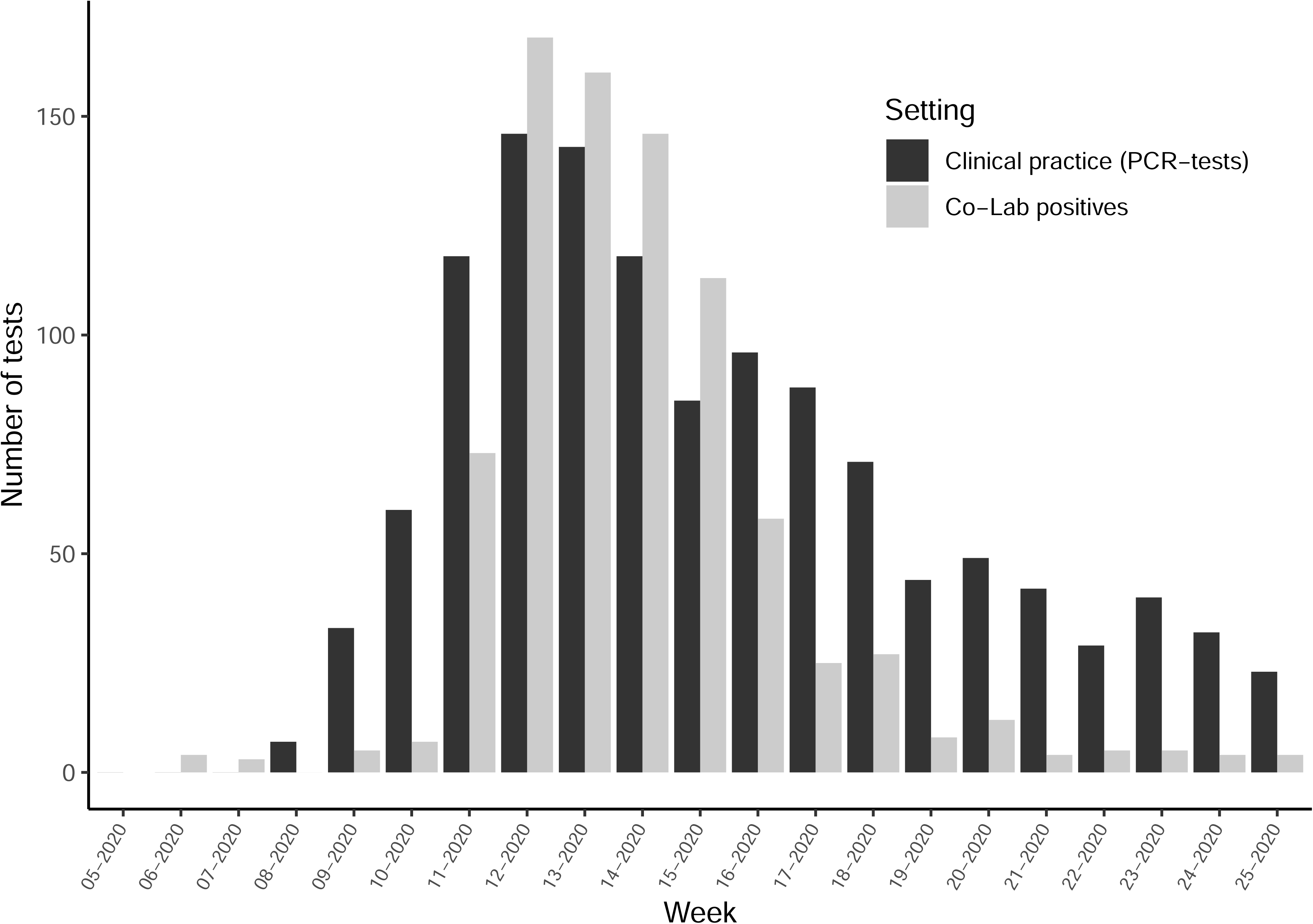
CoLab-guided PCR testing versus routine practice. The black bars represent the number of SARS-CoV-2-PCR tests performed on a weekly basis in the Catharina Hospital. The grey bars represent the number of positives by using the prevalence dependent binary CoLab-classification (in this setting the COVID-19 ED prevalence of the previous week determines the threshold above which a CoLab-score is considered positive in the week thereafter). If PCR testing was performed for all CoLab-positives, the sensitivity would not by lower than a single PCR-test (see **Table 3**) and this would result a reduction in the number of PCR tests.

### Temporal validation

As the CoLab-score was developed in our center after the first COVID-19-wave in the Netherlands, the performance was evaluated in our center during the subsequent second wave. In the Netherlands, the second wave started in August 2020. During this period lab results from 5100 ED presentations were collected. After applying the inclusion flow as shown in **Figure 1B**, 4.333 presentations remained, of which 165 were associated with a COVID-19 infection.

The mean prevalence in this period was 3.8%, which is approximately half of the prevalence during the first period. The AUC of the CoLab-score in the temporal validation set is 0.895 (95% CI: 0.866 to 0.932). The performance in terms of specificity and NPV is similar to the development cohort, whereas the sensitivity and PPV are slightly lower, although 95% CIs overlap (see **Tables 3 and 4**). In terms of calibration, the observed fraction of COVID-19 positives appears slightly higher than expected (**Figure 6A**). When using the prevalence-dependent binary classification, the sensitivity is 0.868 and the NPV is 0.993 (**Table 4**), which is somewhat lower than the development dataset.

**Table 4:** Diagnostic performance of the CoLab-score in the validation dataset (temporal) and three external hospitals. Sensitivities (sens.), specificities (spec.), positive predicting values (PPV), negative predicting values (NPV), true positives (TP), true negatives (TN), false positives (FP) and false negatives (FN) are shown for fixed cut-offs (CoLab-score 0 till ≤ 4) and the prevalence dependent binary CoLab-classification (Pos/Neg). In the last column the fraction of patients at or below the cut-off are shown. For CoLab-score 0, the numbers with round brackets indicate the number of false negatives when only patients with symptom duration >1 and <10 days are taken into account. Note that “0” lists the sensitivity and NPV of CoLab-score 0 and “≤ 4” lists the specificity and PPV of CoLab-score 5.

| CoLab Score | Validation set | Sens. | Spec. | PPV | NPV | TP | TN | FP | FN | % |
| --- | --- | --- | --- | --- | --- | --- | --- | --- | --- | --- |
| 0 | Temporal | 0.958 | 0.410 | 0.060 | 0.996 | 158 | 1708 | 2460 | 7 (2) | 40 |
|  | Center 1 | 1.000 | 0.333 | 0.059 | 1.000 | 52 | 410 | 822 | 0 (0) | 32 |
|  | Center 2 | 0.960 | 0.352 | 0.050 | 0.996 | 95 | 985 | 1815 | 4 (2) | 34 |
|  | Center 3 | 0.973 | 0.322 | 0.131 | 0.991 | 327 | 1034 | 2175 | 9 (5) | 29 |
|  | Combined | 0.969 | 0.363 | 0.080 | 0.995 | 632 | 4137 | 7272 | 20 (9) | 34 |
| ≤1 | Temporal | 0.867 | 0.777 | 0.133 | 0.993 | 143 | 3239 | 929 | 22 | 75 |
|  | Center 1 | 0.923 | 0.696 | 0.113 | 0.995 | 48 | 857 | 375 | 4 | 67 |
|  | Center 2 | 0.929 | 0.680 | 0.093 | 0.996 | 92 | 1905 | 895 | 7 | 66 |
|  | Center 3 | 0.917 | 0.675 | 0.228 | 0.987 | 308 | 2167 | 1042 | 28 | 62 |
|  | Combined | 0.906 | 0.716 | 0.154 | 0.993 | 591 | 8168 | 3241 | 61 | 68 |
| ≤2 | Temporal | 0.776 | 0.885 | 0.211 | 0.990 | 128 | 3688 | 480 | 37 | 86 |
|  | Center 1 | 0.808 | 0.812 | 0.154 | 0.990 | 42 | 1001 | 231 | 10 | 79 |
|  | Center 2 | 0.869 | 0.802 | 0.134 | 0.994 | 86 | 2246 | 554 | 13 | 78 |
|  | Center 3 | 0.893 | 0.795 | 0.313 | 0.986 | 300 | 2552 | 657 | 36 | 73 |
|  | Combined | 0.853 | 0.832 | 0.224 | 0.990 | 556 | 9487 | 1922 | 96 | 79 |
| ≤3 | Temporal | 0.648 | 0.956 | 0.368 | 0.986 | 107 | 3984 | 184 | 58 | 93 |
|  | Center 1 | 0.750 | 0.910 | 0.260 | 0.989 | 39 | 1121 | 111 | 13 | 88 |
|  | Center 2 | 0.687 | 0.899 | 0.193 | 0.988 | 68 | 2516 | 284 | 31 | 88 |
|  | Center 3 | 0.768 | 0.888 | 0.417 | 0.973 | 258 | 2848 | 361 | 78 | 83 |
|  | Combined | 0.724 | 0.918 | 0.334 | 0.983 | 472 | 10469 | 940 | 180 | 88 |
| ≤4 | Temporal | 0.509 | 0.981 | 0.519 | 0.981 | 84 | 4090 | 78 | 81 | 96 |
|  | Center 1 | 0.654 | 0.952 | 0.366 | 0.985 | 34 | 1173 | 59 | 18 | 93 |
|  | Center 2 | 0.556 | 0.953 | 0.294 | 0.984 | 55 | 2668 | 132 | 44 | 94 |
|  | Center 3 | 0.667 | 0.931 | 0.502 | 0.964 | 224 | 2987 | 222 | 112 | 87 |
|  | Combined | 0.609 | 0.957 | 0.447 | 0.977 | 397 | 10918 | 491 | 255 | 93 |
| Pos / Neg | Temporal | 0.865 | 0.867 | 0.217 | 0.993 | 135 | 3189 | 488 | 21 | - |
|  | Center 1 | 0.941 | 0.824 | 0.191 | 0.997 | 48 | 948 | 203 | 3 | - |
|  | Center 2 | 0.857 | 0.861 | 0.193 | 0.994 | 84 | 2179 | 351 | 14 | - |
|  | Center 3 | 0.899 | 0.571 | 0.230 | 0.976 | 295 | 1314 | 988 | 33 | - |
|  | Combined | 0.883 | 0.801 | 0.216 | 0.991 | 567 | 8277 | 2054 | 75 | - |

Based on these findings, we investigated the relation between the CoLab-linear predictor and duration of COVID-19-related symptoms at ED presentation in the *”PCR positive”* presentations of both the internal and temporal validation datasets. As shown in **Figure 4**, the CoLab-linear predictor increases with the duration of symptoms and gradually decreases after day 7. As a consequence, some COVID-19 patients with early or late presentation after onset of symptoms are missed by the CoLab-score. Optimal performance of the CoLab-score is achieved when the onset of symptoms is > 1 and < 10 days prior to ED presentation.

**Figure 4:**
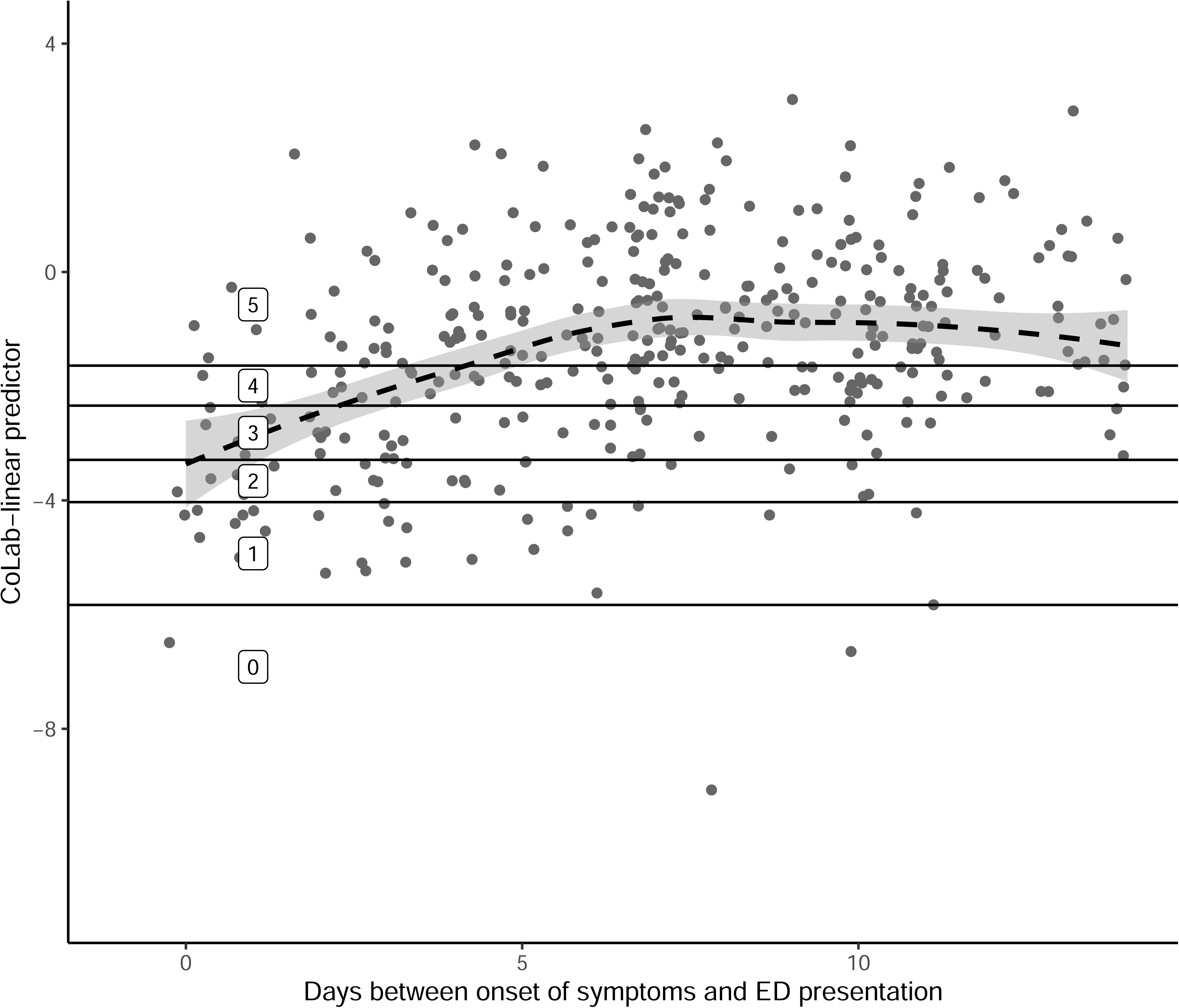
Association between the CoLab-linear predictor and the duration of COVID-19-related symptoms. For all PCR-positive ED presentations in the development and temporal validation dataset, the CoLab-linear predict is plotted against the duration of COVID-related symptoms as registered in the electronic patient records. Patients with unknown duration are not plotted. Patients without symptoms were plotted at 0 days. The solid horizontal lines represent the CoLab-score thresholds, the dashed line is a LOESS regression curve with 95% CI. As the duration of symptoms is an integer, some random jitter was added to the days, for visualization purposes. Note that only the first 14 days are shown in this graph.

### External validation

For external validation, data obtained from three other centers were used. The data were processed analogous to the development dataset. The inclusion flow is summarized in **Figure 5**.

**Figure 5:**
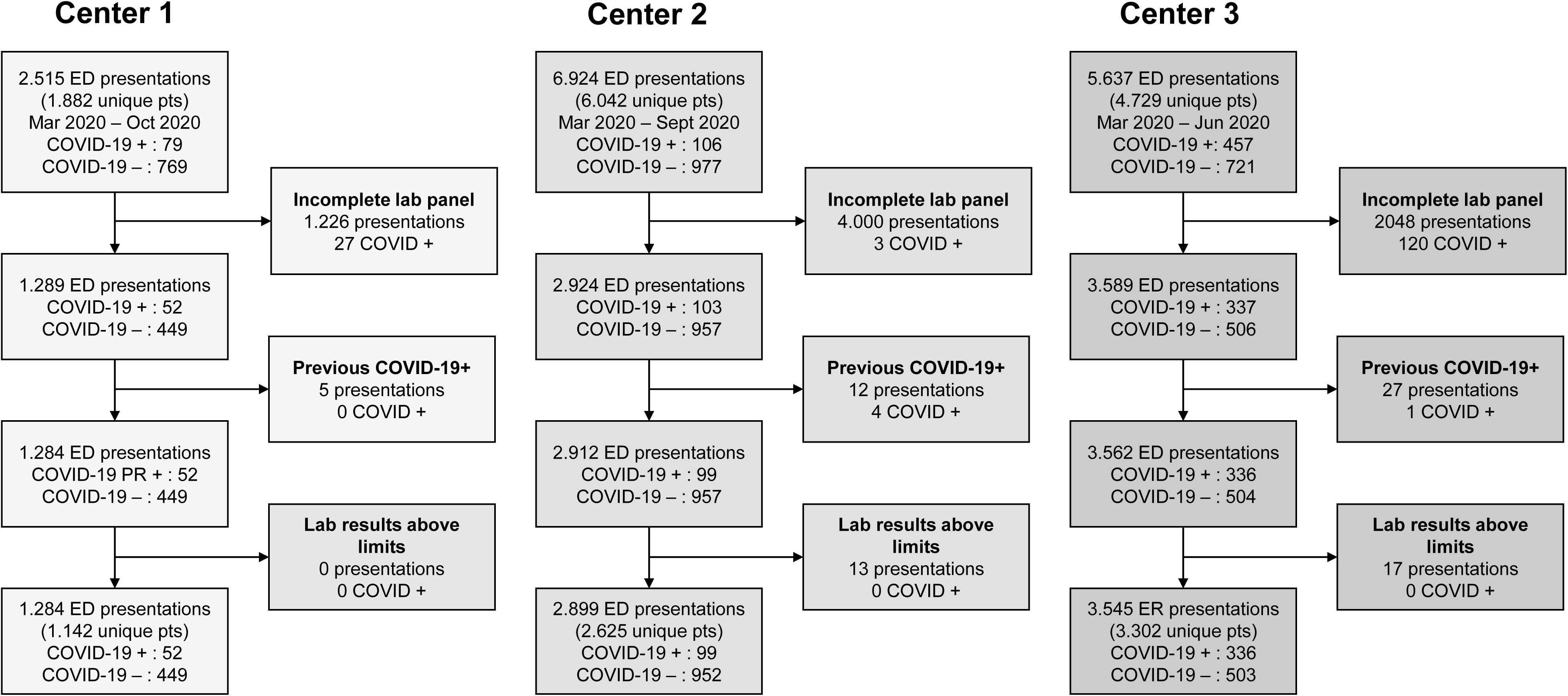
Inclusion flow of ED patients in three external centers. All emergency department (ED) presentations with routine venous blood sampling were included. Missingness of lab panels was assessed for the 11 variables in the CoLab-score (see Table 2). Re-presentations after a positive PCR result or clinical COVID-19 registration were excluded as “previous COVID-19+”. Presentations with any laboratory result above the limits of the CoLab-score (see Table 2) were excluded.

The large number of patients that are excluded in the first step (incomplete laboratory panel) in center 1 and center 2 is due to the fact that only ED presentations for internal medicine were tested for all 10 laboratory tests of the CoLab-score. The ED lab panel of other disciplines (e.g. urology, surgery or pediatrics) differed and did not contain the required tests. Nevertheless, the majority of COVID-19 ED presentations were examined by clinicians of internal medicine, which is reflected by the low number of PCR-positive patients excluded. Other causes of missing lab results were mainly due to assay interference. COVID-19 prevalence differed between the three centers (4.0%, 3.4% and 9.5% respectively) and was lower in centers 1 and 2, and higher in center 3 than the internal validation prevalence. The AUC’s of the CoLab-score are 0.904 (95% CI: 0.866 to 0.942), 0.886 (95% CI: 0.851 - 0.922) and 0.891 (95% CI: 0.872 - 0.909), for centers 1, 2, and 3 respectively.

Performance in terms of sensitivity and specificity is shown in **Table 4**. Also the PPVs and NPVs are shown, although these are largely dependent on prevalence. Although the sensitivity of the prevalence dependent binary CoLab-classification in some centers is somewhat lower than in the development and temporal dataset, the sensitivity of CoLab-score 0 in all centers is ≥ 0.96. Therefore, the NPV of CoLab-score 0 was more than 99%. However, when only presentations were taken into account with duration of symptoms >1 and < 10 days prior to presentation, the NPV for all patients with CoLab-score 0 was >99.8% (**Table 4**).

Calibration plots for external centers are shown in **Figure 6B-D**, the observed fraction of COVID-19 positives is slightly lower than expected in centers 1 and 2. For center 3, low probabilities appear slightly underestimated and high probabilities slightly overestimated.

**Figure 6:**
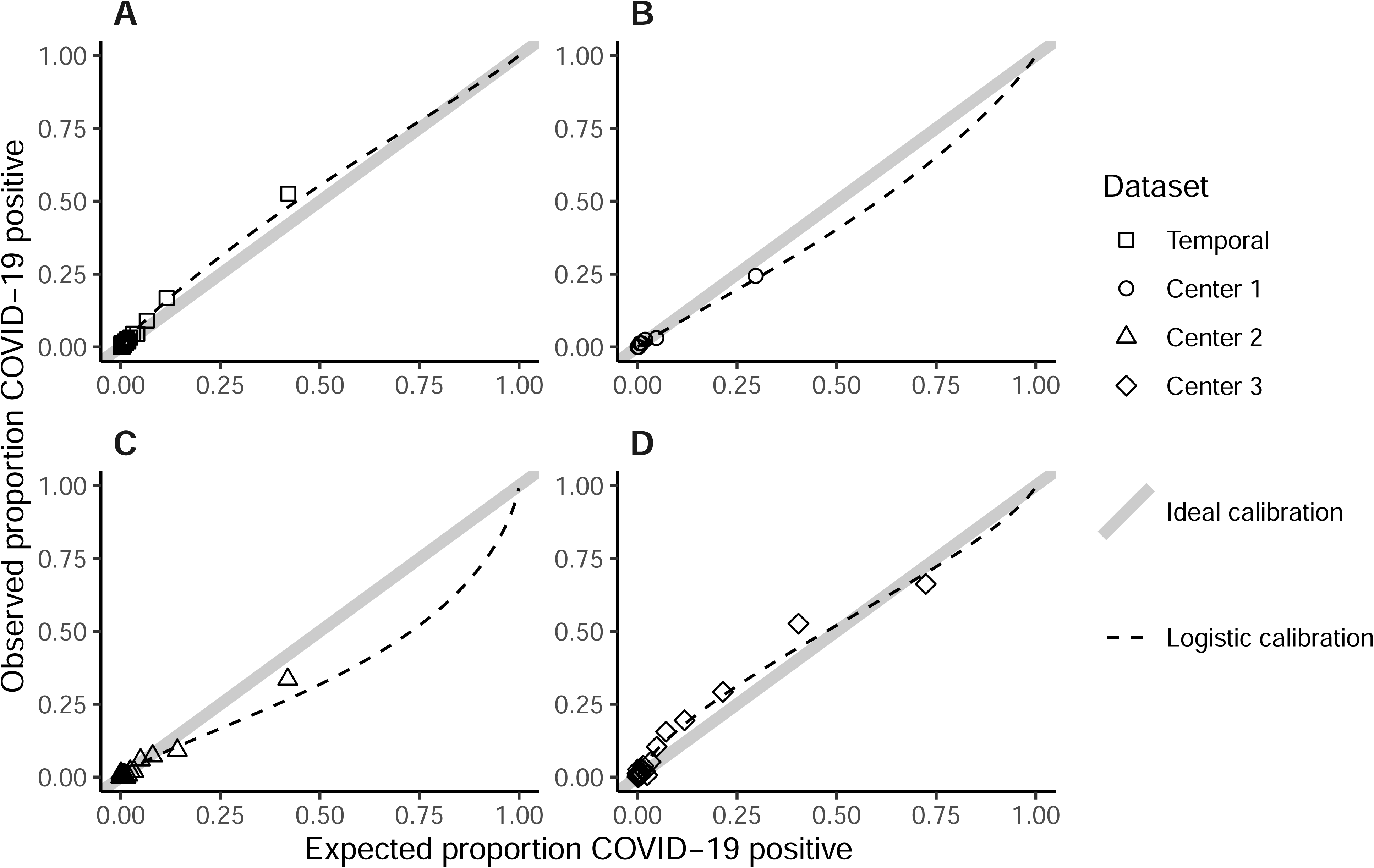
CoLab-score calibration plots of the temporal validation (A), external validation center 1 (B), external validation center 2 (C) and external validation center 3 (D). In the calibration plots, the proportion of observed COVID-19 positives versus expected probabilities are plotted. Observations are grouped with an average of 150 observations per group. The expected probabilities follow from applying the inverse logit function to the CoLab-linear predictor calculated from **Table 2**. If the observed proportion in an external dataset is lower than the expected proportion, this means risks are over-estimated, if the observed fraction is higher, risks are under-estimated. Ideally, observed proportions are equal to expected proportions, this ideal-calibration-line is shown as a straight line through the origin with a slope of 1. The logistic calibration line is a logistic regression fit of the predicted probabilities. [Intercept, slope] for plots A-D: A [0.22, 0.92], B [-0.39, 0.92], C [-0.76, 0.77], D [0.08, 0.79]. Although no validation datasets show perfect calibration, this is the result of differences in COVID-19 prevalence in each center and differences in calibration of laboratory equipment (see Supplemental Figure 6).

**Supplemental Figure 6: Probability density plots of laboratory parameters.**

*Probability density plots are shown for all control patients of the development dataset and the three external centers. Ideally all distributions should overlap since this implies that control patient populations are most likely similar in the development dataset to the external datasets*.

## Discussion

Given the impact of COVID-19 on society and healthcare, there is a need for simple and fast detection of patients with a possible COVID-19 infection in the ED. The CoLab-score described in this study, is a fast and accurate risk score to triage patients presenting at the ED based on routine blood biomarkers. The prevalence-dependent binary CoLab-classification preserves a balance between PCR testing - and not testing, even in situations of changing prevalence. In one third of the ED presentations, PCR testing could be safely omitted with a negative predictive value of 0.995. At low or rapidly decreasing prevalence, the CoLab-classification could save more than half of the PCR tests.

The main strength of this study is that the CoLab-score can be used as a means of early-warning or triaging tool for the entire ED population, regardless of presenting symptoms. This is in contrast to the vast majority of COVID-19 diagnostic models that have been developed on a sub-population of PCR-tested patients (Brinati et al., 2020; Fink et al., 2020; Giamello et al., 2020; Joshi et al., 2020; Kurstjens et al., 2020; Plante et al., 2020; Qin et al., 2020; Tordjman et al., 2020). Moreover, the CoLab-score requires only routine blood tests instead of (features from) imaging such as CT-scans or one or more laboratory test results that are not routinely measured in the ED such as interleukin-6 or 3-hydroxybuteric acid (Wynants et al., 2020). Although this is a retrospective case-control study, there is some unavoidable missing data. In our cohort 13 – 18% of the ED presentations could not be used due to one or more missing laboratory results. This is similar to other studies; 22% (Kurstjens et al., 2020), 17% (Joshi et al., 2020) and 11% (Tordjman et al., 2020). We do not expect that presentations with missing data lead to a severe inclusion bias, but important to note is that the full lab panel was not always requested for pediatric, obstetric and surgery patients. Ideally, probabilities predicted by a model should be well calibrated (Van Calster et al., 2019). However, when the prevalence changes over time, the mean prevalence may not reflect the current prevalence of a center at any given time and calibration is never guaranteed. We chose to categorize the linear predictor into discrete prevalence-based scoring categories. A downside of the CoLab prevalence-dependent binary classification is that false negative results may occur, this is mostly the case when the prevalence increases rapidly. As the threshold depends on the prevalence of the previous week, the threshold is lagging behind the actual prevalence. Estimating the prevalence more frequently during rapidly changing prevalence alleviates this problem but may be more time consuming and prone to stochastic variation. Nevertheless, when absolute certainty to rule-out COVID-19 is required, a CoLab-score of 0 can still be used, unless the prevalence is greater than 40%. In this study we did not investigate whether the CoLab-score might be increased by other virus infections, but in an historic patient cohort we did not find discriminative ability in separating between influenza-PCR-negative and PCR-positive patients (data not shown).

One study that was comparable to our study was published by Plante et. al. (Plante et al., 2020). Their study is similar in terms of outcome and used pre-pandemic patients as controls and PCR positive patients as cases. Also the input variables consisted solely of 15 routinely laboratory tests [4]. Interestingly, their machine learning approach also identified eosinophils, leukocytes, basophils, erythrocytes, albumin and bilirubin as important features. However, one of the other contributing features was bicarbonate, which is generally only requested for a subgroup of ED patients requiring arterial blood gas analysis. This could explain the 63.6% drop outs due to missing data. Furthermore, their model is not easily implemented in current hospital IT systems.

Currently, five hospitals in the Netherlands have already implemented the CoLab-score in daily patient care in the ED, three other centers are in the process of implementation. Main reasons to adopt the CoLab-score, were scarcity in PCR-reagents and long turnaround times of PCR results by centralized PCR-facilities. This underlines the strengths of CoLab-score, i.e. it safely rules out COVID-19 for one third of the ED patients and is instantly reported once the underlying laboratory results are available, usually within one hour.

As we have developed a powerful tool to triage patients at the ED, future research is needed to determine if the CoLab-score also identifies COVID-19 patients with new SARS-CoV-2 mutations. However, since the CoLab-score reflects the biochemical consequences of the virus and not the virus itself, we expect that the CoLab-score is also sensitive for future mutations of SARS-CoV-2.

## Funding

This study was undertaken without any external funding.

## Impact statement

The CoLab-score can rapidly and safely exclude a possible COVID-19-infection in patients presenting at the ER, based on ten routine laboratory tests and age.

## Competing interests

No competing interests declared

## Author contributions

Arjen-Kars Boer: Conceptualization; Resources; Data curation; Formal analysis; Supervision; Validation; Investigation; Visualization; Methodology; Writing - original draft; Writing - review and editing Ruben Deneer: Conceptualization; Data curation; Software; Formal analysis; Validation; Visualization; Methodology; Writing - original draft; Writing - review and editing Maaike Maas: Conceptualization; Supervision; Writing - review and editing Heidi Ammerlaan: Conceptualization; Resources; Supervision; Writing - review and editing Roland van Balkom: Conceptualization; Supervision; Writing - review and editing Mathie Leers: Resources; Data curation; Validation; Writing - review and editing Remy Martens: Resources; Data curation; Validation; Writing - review and editing Madelon Buijs: Resources; Data curation; Validation; Writing - review and editing Jos Kerremans: Resources; Data curation; Validation; Writing - review and editing Muriël Messchaert: Resources; Data curation; Validation; Writing - review and editing Jeroen van Suijlen: Resources; Data curation; Validation; Writing - review and editing Natal van Riel: Resources; Supervision; Methodology; Writing - review and editing Volkher Scharnhorst: Conceptualization; Resources; Supervision; Project administration; Writing - review and editing

## Funding

The funders had no role in study design, data collection and interpretation, or the decision to submit the work for publication.

## Data Availability

Source data to generate figures and tables is provided. It is however, not possible to provide the development dataset used to fit the model since this contains patient data which is subject to EU GDPR regulation. Therefore the source data for Table 1 is split into categories and ordered to prevent patient re-identification. If an interested researcher would like to obtain access to the development dataset, an e-mail to the manager of the data with a research proposal would have to be sent. This proposal will be reviewed by the hospital IRB.

N/A

## Ethics

Human Subjects: Yes Ethics Statement: The study was reviewed by the Medical research Ethics Committees United (MEC-U) under study number W20.071, which confirmed that the Medical Research Involving Human Subjects Act (In Dutch: WMO) does not apply to this study. The study was thereafter reviewed and approved by the internal hospital review board. Clinical Trial: No Animal Subjects: No

## Data Availability

The datasets used to fit the model cannot be released publicly due to the risk of patient re-identification through individual health record data. The authors will consider requests for data sharing as part of an (academic) collaboration after approval by the hospital board of directors and subject to ethical approval and data transfer agreements in accordance with the GDPR regulations. This applies to the development and temporal validation datasets.

## Acknowledgements

Not applicable.

## Notes

### Competing Interest Statement

The authors have declared no competing interest.

### Clinical Trial

The study was reviewed by the Medical research Ethics Committees United (MEC-U) under study number W20.071, which confirmed that the Medical Research Involving Human Subjects Act (In Dutch: WMO) does not apply to this study. The study was thereafter reviewed and approved by the internal hospital review board.

### Funding Statement

This was an investigator-initiated study and no funding was received for this study.

### Author Declarations

Medical research Ethics Committees United (MEC-U) https://www.mec-u.nl/

